# Real-world effectiveness of molnupiravir and nirmatrelvir/ritonavir against mortality, hospitalization, and in-hospital outcomes among community-dwelling, ambulatory COVID-19 patients during the BA.2.2 wave in Hong Kong: an observational study

**DOI:** 10.1101/2022.05.26.22275631

**Authors:** Carlos K.H. Wong, Ivan C.H. Au, Kristy T.K. Lau, Eric H. Y. Lau, Benjamin J. Cowling, Gabriel M. Leung

**Affiliations:** Department of Pharmacology and Pharmacy, LKS Faculty of Medicine, The University of Hong Kong, Hong Kong SAR, China; Department of Family Medicine and Primary Care, LKS Faculty of Medicine, The University of Hong Kong, Hong Kong SAR, China; Laboratory of Data Discovery for Health (D24H), Hong Kong Science and Technology Park, Hong Kong SAR, China; WHO Collaborating Centre for Infectious Disease Epidemiology and Control, School of Public Health, LKS Faculty of Medicine, The University of Hong Kong, Hong Kong SAR, China

## Abstract

**Background:** Evidence evaluating real-world effectiveness of oral antivirals against Omicron variants is lacking.

**Methods:** An unselected, territory-wide cohort of all initially non-hospitalized patients with an officially registered diagnosis of SARS-CoV-2 infection between 26th February and 3rd May 2022 during the Omicron BA.2.2 wave in Hong Kong, was identified. We undertook a retrospective cohort design as primary analysis, and case-control design as sensitivity analysis. Outpatient oral antiviral users were matched with controls using 1:10 propensity-score matching. Study outcomes were mortality, COVID-19-related hospitalization, composite outcome of in-hospital disease progression (in-hospital mortality, invasive mechanical ventilation, or intensive care unit admission) and its individual outcomes. Hazard ratios (HR) were estimated by Cox regression, and odds ratios in oral antiviral users compared with non-users by logistic regression. Subgroup analyses evaluated the associations by vaccination status and age.

**Findings:** Among 1,072,004 non-hospitalized COVID-19 patients, 5,257 and 5,663 were initiated molnupiravir and nirmatrelvir/ritonavir in the community setting with a median follow-up of 42 and 38 days, respectively. Molnupiravir use was associated with lower risks of mortality (HR=0·61, 95%CI=0·46-0·82, p<0·001) and in-hospital composite outcome (HR=0·64, 95%CI=0·50-0·83, p<0·001) than non-use, while that of hospitalization was comparable to controls (HR=1·06, 95%CI=0·97-1·16, p=0·191). Nirmatrelvir/ritonavir use was associated with lower risks of mortality (HR=0·25, 95%CI=0·13-0·47, p<0·001), hospitalization (HR=0·69, 95%CI=0·60-0·79, p<0·001), and in-hospital outcome (HR=0·47, 95%CI=0·31-0·71, p<0·001) than non-use. Similar protective effects of nirmatrelvir/ritonavir were observed across vaccination status (fully vaccinated versus otherwise) and age (dichotomized at 65 years), whereas those for molnupiravir were less consistent. Findings from case-control analysis broadly confirmed those of primary analysis.

**Interpretation:** Amid the Omicron BA.2.2 wave, early initiation of oral antivirals among non-institutionalised COVID-19 patients was associated with reduced risks of mortality and in-hospital outcomes. Nirmatrelvir/ritonavir use was associated with greater and more consistent protection than molnupiravir.

**Funding:** Health and Medical Research Fund, Food and Health Bureau

**Research in context:** *Evidence before this study:* Oral antivirals have been initiating in non-hospitalized COVID-19 patients to lower their risks of hospitalization and death, and hence to reduce the burden on healthcare systems. We searched Scopus and PubMed for studies until 25 May 2022 using the search terms “SARS-CoV-2 OR COVID-19” AND “molnupiravir OR Lagevrio OR EIDD-2801” OR “nirmatrelvir OR Paxlovid OR PF-07321332”. Major studies examining the outpatient use of molnupiravir and nirmatrelvir/ritonavir are MOVe-OUT and EPIC-HR trials, respectively. Both have been conducted among unvaccinated, non-hospitalized patients with mild-to-moderate COVID-19 who are at risk of progression to severe disease, during a pandemic wave of SARS-CoV-2 Delta variant. Early initiation of molnupiravir or nirmatrelvir/ritonavir within five days of symptom onset has been associated with relative risk reduction of hospitalization or death by 30% and 88%, respectively. Considering the real-world evaluation of the two oral antivirals against the currently circulating Omicron variant, only one single-center, retrospective review of solid organ transplant recipients with COVID-19 has been conducted; yet their results are unlikely generalizable to other populations given its specific patient group and small sample size. Real-world effectiveness of oral antivirals is urgently needed to inform their clinical use in COVID-19 patients, considering their vaccination status and the variant of concern.

*Added value of this study:* To the best of our knowledge, this is one of the first real-world studies exploring the clinical use of oral antivirals during a pandemic wave dominated by SARS-CoV-2 Omicron variant. A territory-wide, retrospective cohort study was conducted to examine the effectiveness of molnupiravir and nirmatrelvir/ritonavir in community-dwelling COVID-19 patients. Early initiation of molnupiravir or nirmatrelvir/ritonavir within five days of symptom onset was associated with significant reduction of all-cause mortality risk by 39% and 75%, respectively, compared to not using any oral antivirals. Nirmatrelvir/ritonavir use was also associated with a reduced risk of COVID-19-related hospitalization by 31%, which was consistently observed across age and vaccination status. In terms of disease progression, both oral antivirals were effective in lowering the risk of in-hospital death, which was again more substantial with nirmatrelvir/ritonavir than molnupiravir. Intriguingly, the need for invasive ventilation might be reduced among molnupiravir users compared to matched controls.

*Implications of all the available evidence:* Based on relative efficacy, our findings give support to current guidelines prioritizing nirmatrelvir/ritonavir use over molnupiravir in community-dwelling COVID-19 patients who are at high risk of hospitalization or progression to severe disease, should the former be accessible and clinically appropriate. Amid a pandemic wave of the Omicron variant, real-world effectiveness of oral antivirals in reducing the mortality risk of community-dwelling COVID-19 patients has been demonstrated in this study consisting mostly of the elderly and those who had not been fully vaccinated, extending beyond the evidence demonstrated in clinical trials among those of the Delta variant and who were at risk of severe COVID-19 from being overweight/obese. Several clinical trials (namely RECOVERY and PANORAMIC) and observational studies of the two oral antivirals are ongoing, and further research is needed to confirm our results in other patient populations and healthcare settings.

## Introduction

Oral antiviral drugs including molnupiravir (Lagevrio) and nirmatrelvir/ritonavir (Paxlovid) are novel options for treating adult patients with coronavirus disease 2019 (COVID-19) that have been shown in clinical trials of unvaccinated patients prior to the Omicron phase of the ongoing pandemic to reduce hospitalization and mortality.^1-3^ There remains a dearth of evidence from the field under contemporary real-world conditions.^1,4^

Clinical trials including the MOVe-OUT study have shown that early administration of molnupiravir to non-hospitalized patients with mild-to-moderate COVID-19 accelerates viral clearance, alongside a modest relative risk reduction of hospitalization or mortality by 30%.^3,5^

According to the EPIC-HR trial, nirmatrelvir/ritonavir could significantly reduce the rates of hospitalization and mortality by 89% when the drug was initiated within three days of symptom onset, and consistently by 88% within five days of symptom onset, in non-hospitalized patients with mild-to-moderate COVID-19 who were at risk of progression to severe disease.^6^

While both novel oral antivirals have been approved for treatment of mild-to-moderate COVID-19, there is as yet no observational evidence evaluating their real-world effectiveness. In particular, the original trials were carried out mostly during the Delta wave whereas effectiveness against Omicron and its subvariants can only be inferred from laboratory studies so far.^7-10^ Here we assessed the clinical effectiveness of molnupiravir and nirmatrelvir/ritonavir among community-dwelling COVID-19 outpatients in Hong Kong during the Omicron BA.2.2 wave in January to May 2022.

## Methods

### Study design and data sources

A territory-wide observational study was conducted to examine the effectiveness of oral antiviral treatment against COVID-19 mortality or hospitalization in non-institutionalized, aged 18 or above, COVID-19 patients in Hong Kong Special Administrative Region, China, during the Omicron BA.2 wave from 1st January 2022 onward.

We analyzed the electronic medical records of COVID-19 patients (defined by laboratory-confirmed positive of reverse transcription-polymerase chain reaction test, or positive rapid antigen test [RAT]) from the Hospital Authority (HA), a statutory provider of public inpatient services and primary public outpatient services in Hong Kong. Electronic medical records, which included demographic characteristics, date of registered death, hospital admission, emergency department visits, diagnoses, prescription and drug dispensing records, procedures, and laboratory tests, had linkage to anonymized COVID-19 vaccination records provided by the Department of Health using the unique identification numbers. The database has been widely used for high-quality studies to evaluate the effectiveness of drug treatments for COVID-19 at a population level.^11^

Patients who had COVID-19 diagnosis from 26th February 2022 (first oral antiviral drug prescription date) to 3rd May 2022, were eligible. Patients who were aged below 18, and admitted to hospital before the COVID-19 diagnosis, death on or before the COVID-19 diagnosis, or residents at the residential care homes for the elderly (RCHE), were excluded.

We conducted the retrospective cohort study design as primary analysis, and case-control study design as sensitivity analysis for internal validation. The index date within the cohort study was defined as the date of oral antiviral initiation in treatment cohort, and the date of COVID-19 diagnosis in control cohort. Control cohort was selected from the patients with COVID-19 diagnosis prior to admission, and those who did not receive oral antiviral in outpatient setting during the observational period, using the propensity-score in a ratio of 1:10 (described in statistical analyses section below). Patients were observed from the index date to death, outcome events occurrence, crossover of oral antiviral treatment, or the end of observational period (3rd May 2022), whichever came first.

Within the case-control study design, patients with COVID-19 receiving molnupiravir or nirmatrelvir/ritonavir treatment in outpatient setting before the reference date, which was the date of outcome events for cases and 28 days after the COVID-19 diagnosis for controls. Up to ten control patients were randomly matched with each of the case according to age (within the same year), sex, date of COVID diagnosis (within the same date), Charlson Comorbidity Index (CCI), and full SARS-CoV-2 vaccination (with at least two doses of Comirnaty or three doses of CoronaVac).

Our study followed the STROBE (Strengthening the Reporting of Observational Studies in Epidemiology) guidelines. This study was approved by the institutional review board of the University of Hong Kong / Hospital Authority Hong Kong West Cluster (reference no. UW 20-493). Given the extraordinary nature of the COVID-19 pandemic, individual patient-informed consent was not required for this retrospective cohort study using anonymized data.

### Outcome definition

Outcomes of cohort study were 1) all-cause mortality, 2) COVID-19-related hospitalization, 3) a composite in-hospital disease progression outcome (in-hospital mortality, invasive mechanical ventilation [IMV], or intensive care unit admission), and 4) individual in-hospital outcomes.

Case-control study measured the same first three outcomes. For the outcome of all-cause death, we defined cases and controls as patients who died and did not die within 28 days after the COVID-19 diagnosis during the observation period, respectively. For the outcome of COVID-19-related hospitalization, cases were defined as patients admitted to the hospital within 28 days after the COVID-19 diagnosis whilst controls were defined as those without hospital admission within 28 days after the diagnosis. Only the first hospital admission after the diagnosis were used if a patient had multiple hospital admissions. Composite in-hospital outcome applied similar case and control definition.

### Statistical Analyses

In the retrospective cohort design, propensity score models conditional on age, sex, date of COVID-19 diagnosis, CCI and vaccination status in a logistic regression model was performed. Standardized mean differences (SMD) of each covariate between the groups before and after the propensity matching were calculated, and were interpreted as balance when the SMD was below the threshold of 0·1.^12^ Hazard ratios (HR) with 95% confidence intervals (CI) of each outcome between oral antiviral users and their respective matched non-users were estimated using Cox regression models.

In the case-control design as sensitivity analysis, conditional logistic regression was used to examine the association of receiving oral antiviral drug treatment with hospitalization and all-cause mortality among COVID-19 patients. Odds ratios (ORs) in molnupiravir users and nirmatrelvir/ritonavir users compared with non-users were estimated. Subgroup analyses evaluated the associations by vaccination status (fully vaccinated versus not fully vaccinated) and age groups (≤65 versus >65 years). Interactions between oral antiviral drug treatment and subgroups were evaluated.

All statistical analyses were performed using Stata version 17 (StataCorp LP, College Station, TX). All significance tests were two-tailed, where P-value <0·05 was considered statistically significant.

## Results

During the Omicron BA.2.2 wave in Hong Kong, 1,072,004 patients with COVID-19 diagnosis were identified in our study period, where 10,920 of them were initiated on either of the two novel oral antivirals in the community setting (molnupiravir: 5,257; nirmatrelvir/ritonavir: 5,663) (Figure 1). In the retrospective cohort, there were 4,875 molnupiravir users and 48,409 matched controls, in addition to 5,366 nirmatrelvir/ritonavir users and 53,289 matched controls. Demographic and clinical characteristics of non-hospitalized COVID-19 patients are presented in Supplementary Table 1 by oral antiviral use. After matching, patient characteristics were balanced between oral antiviral and respective control groups at baseline, with all SMDs <0·1 (Table 1). Overall, less than half of the cohort had been fully vaccinated. Compared to molnupiravir, the proportion of elderly patients (aged >65 years) was lower in the nirmatrelvir/ritonavir group, as well as that of pre-existing comorbidities. Both oral antivirals were initiated in community-dwelling COVID-19 patients after a median of 2 (interquartile range: 1-4) days since symptom onset.

**Figure 1.**
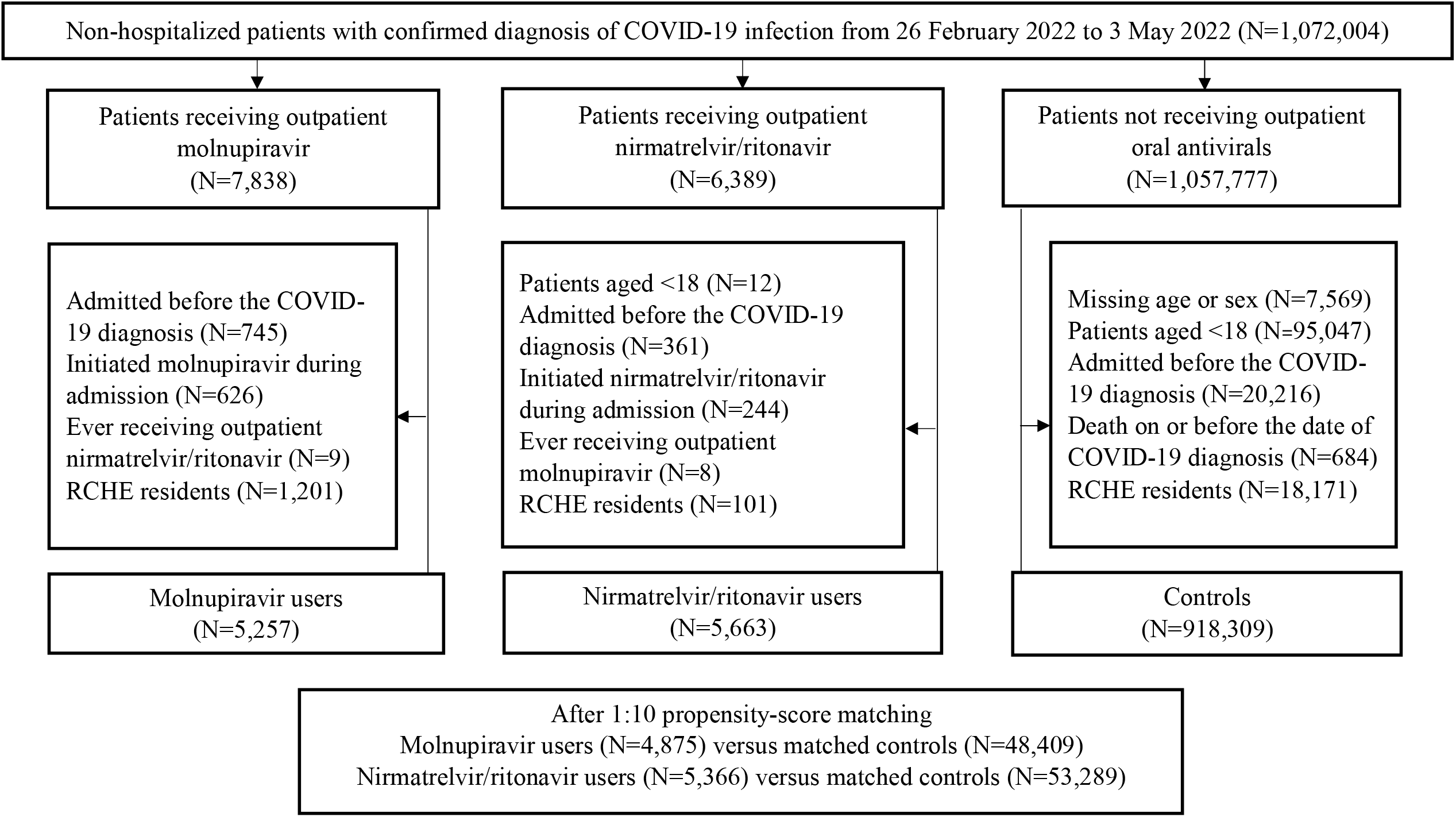
Identification of outpatient molnupiravir users, nirmatrelvir/ritonavir users, and their matched controls among non-hospitalized patients with COVID-19 from 26 February 2022 to 3 May 2022 in Hong Kong Note: RCHE = residential care homes for the elderly

**Table 1.** Baseline characteristics of non-hospitalized patients with COVID-19 in (a) molnupiravir and respective matched control groups, and (b) nirmatrelvir/ritonavir and respective matched control groups after 1:10 propensity score matching

| Baseline characteristics | After 1:10 matching |  |  |  |  |  |  |  |  |  |
| --- | --- | --- | --- | --- | --- | --- | --- | --- | --- | --- |
|  | Molnupiravir<br>(n=4,875) |  | Control<br>(n=48,409) |  | SMD | Nirmatrelvir/ritonavir<br>(n=5,366) |  | Control<br>(n=53,289) |  | SMD |
|  | N / Mean | % / SD | N / Mean | % / SD |  | N / Mean | % / SD | N / Mean | % / SD |  |
| Age, years † | 74.1 | 12.5 | 74.9 | 12.2 | 0.07 | 70.0 | 11.9 | 70.4 | 14.5 | 0.03 |
| 18-40 | 88 | (1.8%) | 423 | (0.9%) |  | 135 | (2.5%) | 1,921 | (3.6%) |  |
| 40-65 | 919 | (18.9%) | 9,855 | (20.4%) | 0.09 | 1,558 | (29.0%) | 16,163 | (30.3%) | 0.07 |
| >65 | 3,868 | (79.3%) | 38,131 | (78.8%) |  | 3,673 | (68.5%) | 35,205 | (66.1%) |  |
| Sex |  |  |  |  | 0.00 |  |  |  |  | 0.01 |
| Male | 2,324 | (47.7%) | 23,070 | (47.7%) |  | 2,480 | (46.2%) | 24,449 | (45.9%) |  |
| Female | 2,551 | (52.3%) | 25,339 | (52.3%) |  | 2,886 | (53.8%) | 28,840 | (54.1%) |  |
| Pre-existing comorbidities |  |  |  |  |  |  |  |  |  |  |
| Charlson's Index † | 3.8 | 1.3 | 3.8 | 1.2 | 0.01 | 3.4 | 1.1 | 3.3 | 1.2 | 0.04 |
| 0-4 | 4,337 | (89.0%) | 44,169 | (91.2%) |  | 5,124 | (95.5%) | 50,861 | (95.4%) |  |
| 5-6 | 355 | (7.3%) | 2,740 | (5.7%) | 0.08 | 200 | (3.7%) | 1,969 | (3.7%) | 0.01 |
| 7-14 | 183 | (3.8%) | 1,500 | (3.1%) |  | 42 | (0.8%) | 459 | (0.9%) |  |
| Fully vaccinated* | 839 | (17.2%) | 7,513 | (15.5%) | 0.05 | 1,896 | (35.3%) | 18,806 | (35.3%) | 0.00 |

Notes: SD = standard deviation; SMD = standardized mean difference
\* Fully vaccinated patients were defined as those with at least 2 doses of Comirnaty or 3 doses of CoronaVac.
† Age and Charlson Comorbidity Index are presented in mean ± SD.

Overall, molnupiravir and nirmatrelvir/ritonavir users were followed for a median of 42 (interquartile range: 36-47) and 38 (interquartile range: 31-42) days, respectively. The cumulative incidences of all-cause mortality, COVID-19-related hospitalization, and in-hospital disease progression between oral antiviral and respective control groups are illustrated in Figure 2. The crude incidence rates of all-cause mortality were 24·2 and 38·0 per 100,000 person-days among molnupiravir users and matched controls, and 5·2 and 20·1 per 100,000 person-days among nirmatrelvir/ritonavir users and their matched controls (Table 2). Molnupiravir use was associated with a significantly lower risk of all-cause mortality than non-use (HR=0·61, 95%CI=0·46-0·82, p<0·001), while that of hospitalization was comparable to control (HR=1·06, 95%CI=0·97-1·16, p=0·191). In contrast, nirmatrelvir/ritonavir use was associated with significantly lower risks of both all-cause mortality (HR=0·25, 95%CI=0·13-0·47, p<0·001) and hospitalization (HR=0·69, 95%CI=0·60-0·79, p<0·001) than non-use.

**Table 2.**
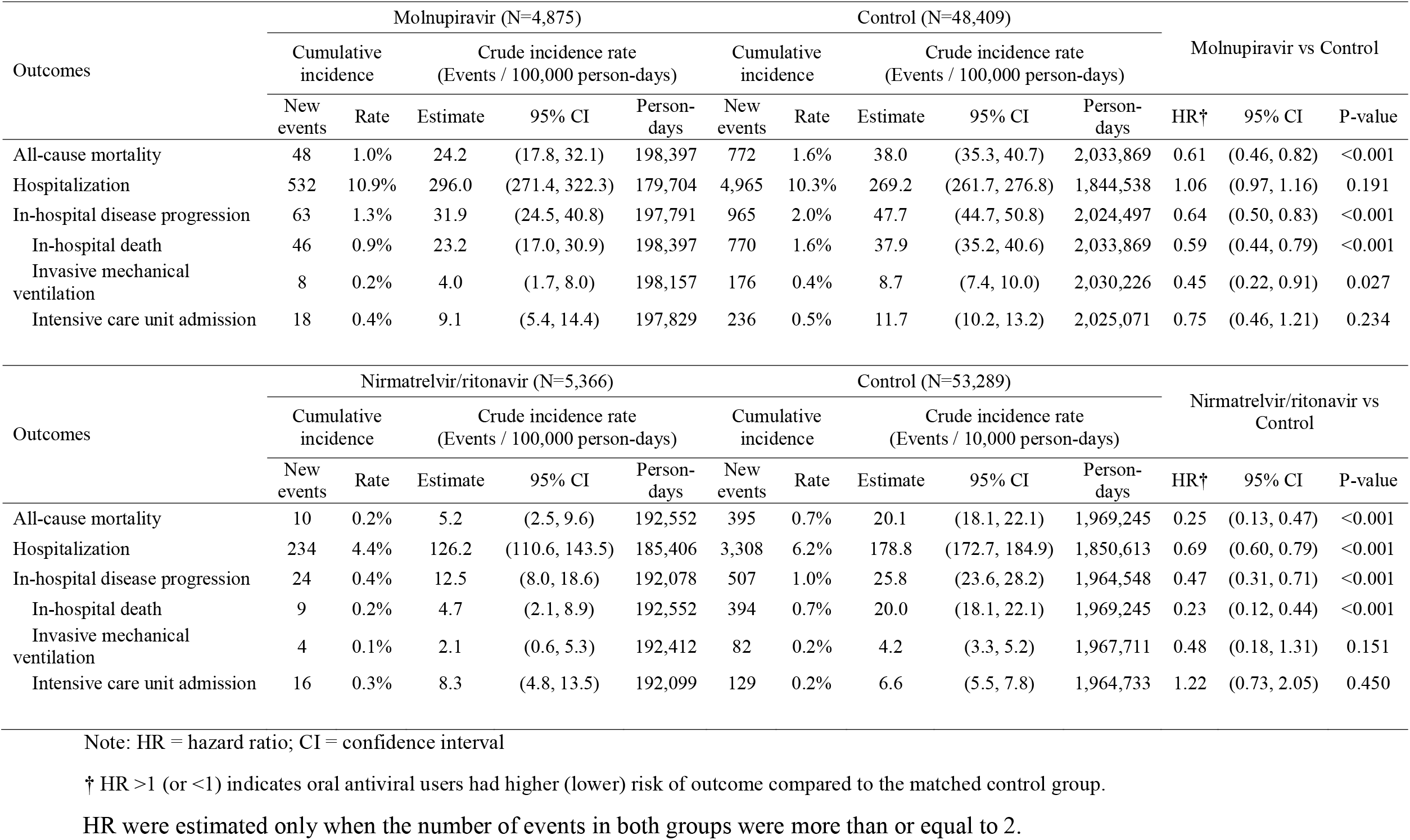
Hazard ratios of outcomes for (a) outpatient molnupiravir users versus matched controls, and (b) outpatient nirmatrelvir/ritonavir users versus matched controls

**Figure 2.**
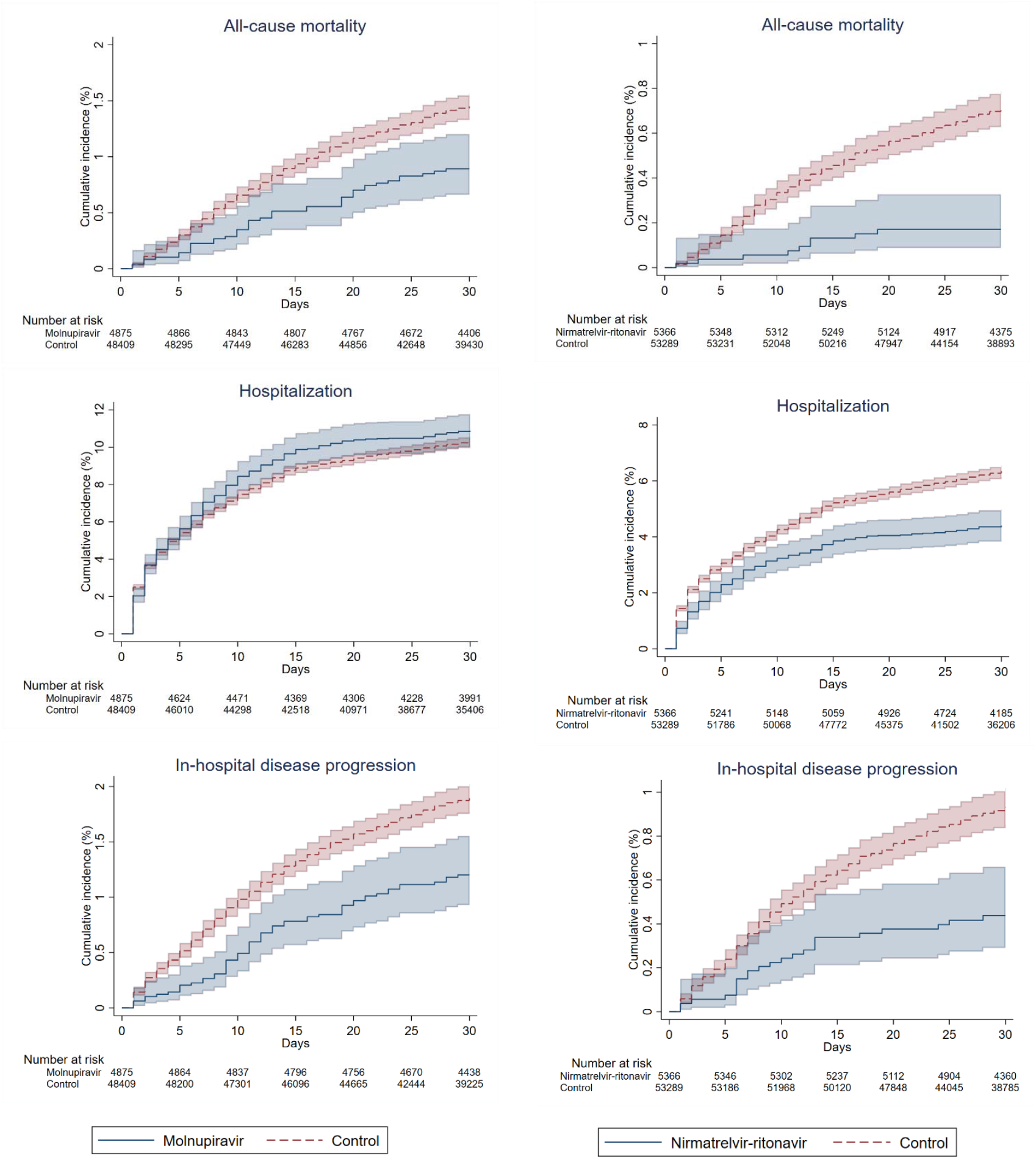
Cumulative incidence plots of (a) all-cause mortality, (b) hospitalization, and (c) in-hospital disease progression for outpatient molnupiravir users versus their matched controls, and (a) all-cause mortality, (b) hospitalization, and (c) in-hospital disease progression for outpatient nirmatrelvir/ritonavir users versus their matched controls

Concerning the in-hospital composite outcome, molnupiravir use was associated with a significantly lower risk of disease progression than non-use (HR=0·64, 95%CI=0·50-0·83, p<0·001), which was consistently observed for its individual outcomes of in-hospital death (HR=0·59, 95%CI=0·44-0·79, p<0·001) and IMV initiation (HR=0·45, 95%CI=0·22-0·91, p=0·027) (Table 2). Risk of ICU admission was comparable between molnupiravir and control groups (HR=0·75, 95%CI=0·46-1·21, p=0·234). On the other hand, while the risk of in-hospital composite outcome was also significantly reduced with nirmatrelvir/ritonavir use than non-use (HR=0·47, 95%CI=0·31-0·71, p<0·001), it was mainly driven by a substantial mortality benefit (HR=0·23, 95%CI=0·12-0·44, p<0·001) than reducing IMV initiation (HR=0·48, 95%CI=0·18-1·31, p=0·151) or ICU admission (HR=1·22, 95%CI=0·73-2·05, p=0·450). The proportion of community-dwelling COVID-19 patients who were admitted, on respiratory support, discharged alive, or dead over the follow-up period are represented in Figure 3 by oral antiviral use.

**Figure 3.**
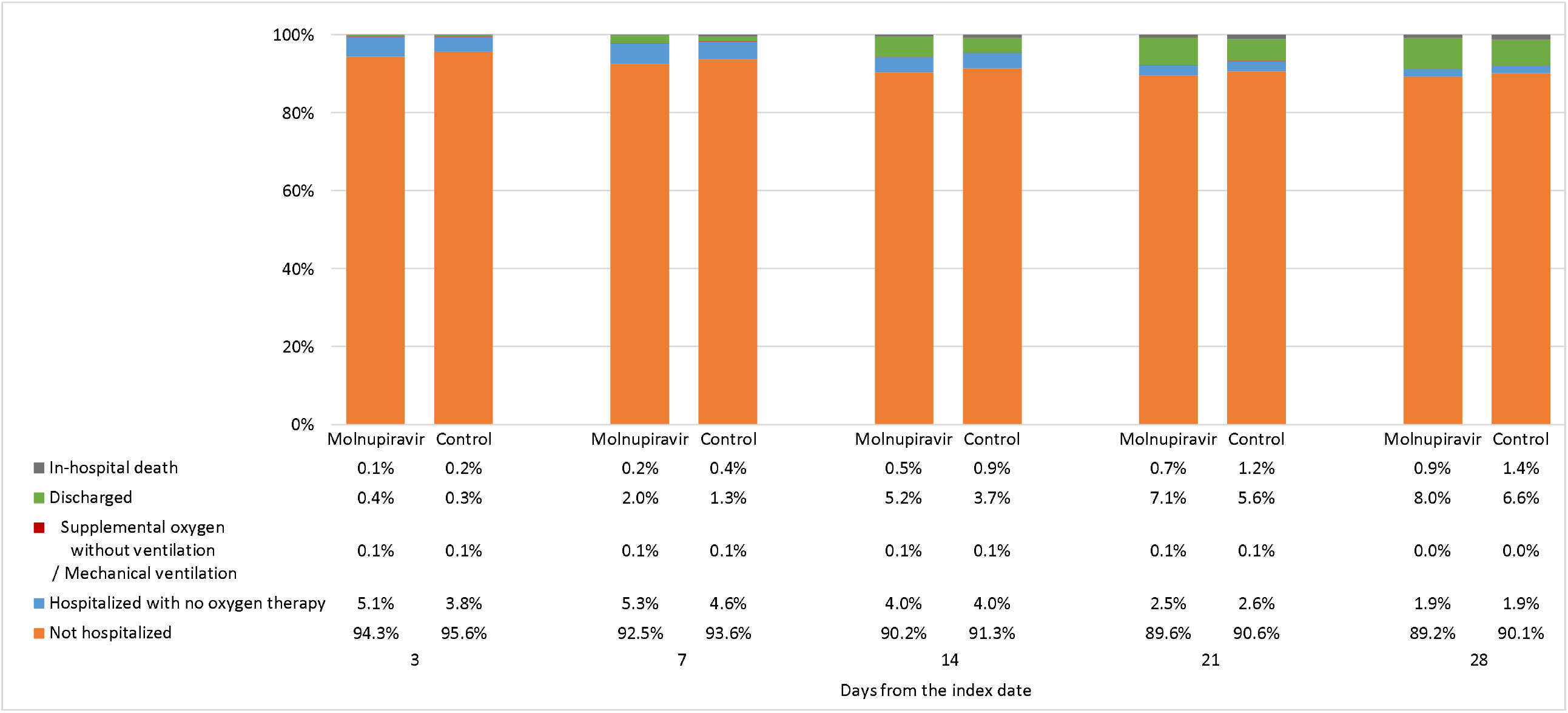
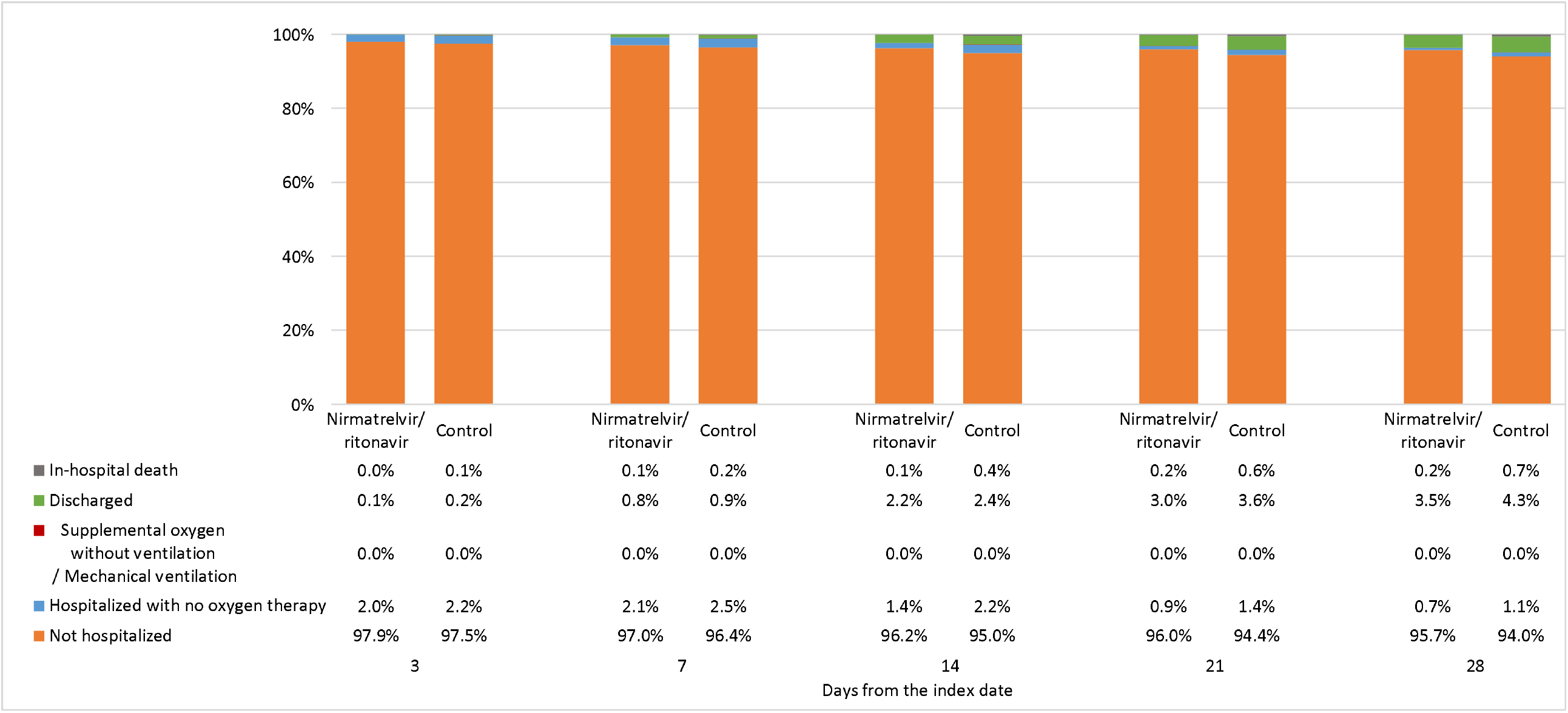
Comparison of disease status at days 3, 7, 14, 21, and 28 after the index date (COVID diagnosis) a) between outpatient molnupiravir users and their matched controls, and b) between outpatient nirmatrelvir/ritonavir users and their matched controls

With respect to the sensitivity analysis using case-control design, patient characteristics in case and control groups for each outcome after matching are reported in Supplementary Table 2. Oral antiviral use was associated with significantly lower odds of all-cause mortality (molnupiravir: OR=0·50, 95%CI=0·36-0·68, p<0·001; nirmatrelvir/ritonavir: OR=0·15, 95%CI=0·06-0·36, p<0·001) and in-hospital disease progression (molnupiravir: OR=0·56, 95%CI=0·41-0·77, p<0·001; nirmatrelvir/ritonavir: OR=0·51, 95%CI=0·31-0·84, p=0·008) than non-use among community-dwelling COVID-19 patients (Table 3). Meanwhile, the lower odds of hospitalization were only significant with nirmatrelvir/ritonavir use (OR=0·54, 95%CI=0·38-0·76, p<0·001), but not molnupiravir (OR=1·01, 95%CI=0·78-1·32, p=0·913). These findings were generally in line with those of the primary analysis.

**Table 3.** Odds ratios of outpatient molnupiravir and nirmatrelvir/ritonavir exposure between cases and controls after matching in case-control study design

| All-cause mortality | Case (N=1,084) | Control (N=10,454) | OR | 95% CI | P-value |
| --- | --- | --- | --- | --- | --- |
| Non-users | 1,038 (95.8%) | 9,407 (90.0%) |  | (reference) |  |
| Molnupiravir users | 41 (3.8%) | 749 (7.2%) | 0.50 | (0.36, 0.68) | <0.001 |
| Nirmatrelvir/ritonavir users | 5 (0.5%) | 302 (2.9%) | 0.15 | (0.06, 0.36) | <0.001 |
| Hospitalization | Case (N=2,052) | Control (N=19,099) | OR | 95% CI | P-value |
| Non-users | 1,955 (95.3%) | 17,950 (94.0%) |  | (reference) |  |
| Molnupiravir users | 63 (3.1%) | 570 (3.0%) | 1.01 | (0.78, 1.32) | 0.913 |
| Nirmatrelvir/ritonavir users | 34 (1.7%) | 581 (3.0%) | 0.54 | (0.38, 0.76) | <0.001 |
| In-hospital disease progression | Case (N=880) | Control (N=8,438) | OR | 95% CI | P-value |
| Non-users | 820 (93.2%) | 7,443 (88.2%) |  | (reference) |  |
| Molnupiravir users | 43 (4.9%) | 698 (8.3%) | 0.56 | (0.41, 0.77) | <0.001 |
| Nirmatrelvir/ritonavir users | 17 (1.9%) | 302 (3.6%) | 0.51 | (0.31, 0.84) | 0.008 |

Notes: OR = Odds ratio; CI = confidence interval

In the subgroup analyses of study outcomes stratified by age and vaccination status (Supplementary Table 3), results comparing molnupiravir and non-use were similar to those of the primary analysis, except for COVID-19-related hospitalization. Molnupiravir use was associated with a significantly lower risk of hospitalization among those who had been fully vaccinated (HR=0·62, 95%CI=0·44-0·89, p=0·009), but not for those who had not completed two-dose vaccine schedule (HR=1·12, 95%CI=1·02-1·23, p=0·013) (p-value for interaction=0·003). Interestingly, molnupiravir use was also associated with a significantly higher risk of hospitalization among younger patients aged ≤65 years (HR=1·59, 95%CI=1·28-1·99, p<0·001), which was not evident in those aged >65 years (HR=0·99, 95%CI=0·89-1·09, p=0·782) (p-value for interaction<0·001). Meanwhile, results comparing nirmatrelvir/ritonavir and non-use were consistent across subgroups, in particularly for the significantly lower risk of COVID-19-related hospitalization that was evident among nirmatrelvir/ritonavir users regardless of age and vaccination status.

## Discussion

In this retrospective cohort of community-dwelling COVID-19 patients during a pandemic wave of SARS-CoV-2 Omicron variant, early initiation of molnupiravir or nirmatrelvir/ritonavir at a median of two days since symptom onset was associated with significant reduction of all-cause mortality risk by 39% and 75%, respectively, compared to not using any oral antivirals.

Nirmatrelvir/ritonavir use was also associated with 31% reduced risk of COVID-19-related hospitalization. In terms of disease progression, both oral antivirals were effective in lowering the risk of in-hospital death, which was again more substantial with nirmatrelvir/ritonavir than molnupiravir. Intriguingly, the need for IMV might be reduced among molnupiravir users compared to matched controls, should the clinical condition of these COVID-19 patients deteriorate and hospitalization is required. In contrast to the reduced risk of hospitalization that was consistently observed among nirmatrelvir/ritonavir users regardless of their age and vaccination status, this was only evident in molnupiravir users who had been fully vaccinated.

Among non-hospitalized patients with mild-to-moderate COVID-19 who were unvaccinated and at risk of progression to severe disease, early initiation of molnupiravir within five days of symptom onset contributed to a relative reduction of hospitalization or death risk by 30% in the MOVe-OUT trial (89% risk reduction for all-cause mortality), and nirmatrelvir/ritonavir by 88% in the EPIC-HR trial.^3,6^ The interim analysis of EPIC-SR trial involving unvaccinated adults at standard risk or vaccinated individuals with at least one risk factor demonstrated a reduction of hospitalization risk by 70% with nirmatrelvir/ritonavir use in non-hospitalized COVID-19 patients, which was of borderline statistical significance (p=0·051).^13-15^ While both oral antivirals have been shown to exhibit robust activity in lowering the viral load substantially relative to placebo,^3,6,14^ the number needed to treat (NNT) was higher with molnupiravir than nirmatrelvir/ritonavir use based on the final results of MOVe-OUT and EPIC-HR trials.^16-18^ Notably, both estimated NNT are expected to increase further in the current wave dominated by Omicron variant, which results in less hospitalization and death compared to Delta variant that circulated during previous trials,^19,20^ and the increasing proportion of individuals who have been fully vaccinated.^15^ Therefore, real-world effectiveness of oral antivirals should be evaluated in specific contexts considering SARS-CoV-2 VOC and population immunity.

At the time of writing, the antiviral effectiveness of molnupiravir and nirmatrelvir/ritonavir against hospitalization among patients infected with SARS-CoV-2 Omicron variant has not been fully evaluated in any published clinical studies. There is only one single-center, retrospective review of outpatient therapies for solid organ transplant recipients with COVID-19; yet their results might not be generalizable to other populations given its specific patient group and small sample size (49 patients receiving molnupiravir, and only one on nirmatrelvir/ritonavir).^21^ Relying on evidence derived from experimental studies, viral replication of Omicron variant is inhibited in treated cell lines, and infection restricted in hamsters treated with oral antivirals.^7- 10,22,23^ It is postulated that Omicron is similarly sensitive to both drugs as previous variants, given most mutations of the Omicron variant are located around the spike protein, and those involving the target enzymes (RdRp and M^pro^, respectively) are distant from their active sites.^7-9,22,23^ As illustrated in the current study conducted amid a pandemic wave of Omicron BA.2 variant, early initiation of nirmatrelvir/ritonavir (within five days of symptom onset) in community-dwelling COVID-19 patients was associated with significant reduction in the risks of all-cause mortality by 75% and hospitalization by 31% compared to non-use, whereas a more modest effect was observed with molnupiravir in lowering the mortality risk by 39%. While both oral antivirals appeared effective in lowering the risks of disease progression and in-hospital death, molnupiravir might offer an additional benefit in reducing the need for invasive ventilation, which will have to be confirmed in further studies. Notably, the interpretation of our results and comparative effectiveness of the two oral antivirals should also take into consideration the potential differences in patient characteristics at baseline, where the proportion of elderly patients (aged >65 years) was higher, and that of fully vaccinated individuals was lower, among molnupiravir users and matched controls than their counterparts on nirmatrelvir/ritonavir. These might have partially contributed to the relatively inferior clinical outcomes with molnupiravir than nirmatrelvir/ritonavir use in this study.

To the best of our knowledge, this is one of the first clinical studies exploring real-world effectiveness of the two oral antivirals in non-hospitalized patients infected with the Omicron variant. Based on relative efficacy, our findings give support to current guidelines prioritizing nirmatrelvir/ritonavir use over molnupiravir in community-dwelling COVID-19 patients who are at high risk of hospitalization or progression to severe disease (as indicated by the old age and incomplete vaccination status of our patients), should the former be accessible and clinically appropriate.^24-27^ While our results suggest similar trends to MOVe-OUT and EPIC-HR trials, discrepancies in the respective effect size may be attributed to differences in the risk profile of patients (with overweight/obesity being the major risk factor of patients in the two clinical trials, and <20% of them were over 60 or 65 years old), and/or the circulating VOC (Omicron in this study versus Delta in previous trials).^3,6^ Furthermore, our subgroup analyses reinforced the significant benefit of nirmatrelvir/ritonavir use in lowering the risk of COVID-19-related hospitalization regardless of age and vaccination status, which could only be observed in molnupiravir users who had been fully vaccinated. Nevertheless, both oral antivirals were effective in reducing the mortality risk of elderly patients or those who had not been fully vaccinated in the community setting.

Regarding the observed differences in relative efficacy between the two oral antivirals, an experimental study has recently hypothesized that drug concentrations in the lungs of COVID-19 patients might play a role.^28^ While remdesivir and EIDD-1931 (active metabolite of molnupiravir) have been identified as substrates of human equilibrative nucleoside transporters (ENT) 1 and 2, such interaction is not evident with nirmatrelvir.^28,29^ As ENT expression and function are likely repressed during COVID-19-induced acute lung injury and tissue hypoxia, pulmonary concentrations of remdesivir and EIDD-1931 may be lower than that of nirmatrelvir in COVID-19 patients, hence higher effectiveness of nirmatrelvir/ritonavir than remdesivir and molnupiravir with controversial findings.^28,30^ In fact, the MOVe-OUT trial has been criticized for its premature termination, imbalances of patient characteristics between treatment and control groups at baseline, inconsistency in the results between interim and final analyses that could not be fully explained by differences in patient characteristics, and the dubious value of molnupiravir given its modest reduction in hospitalization or death rate that failed to reach statistical significance (HR=0·69, 95%CI=0·48-1·01).^3,30-33^ Accordingly, further studies are needed to confirm the real-world effectiveness of molnupiravir and nirmatrelvir/ritonavir in different healthcare settings and COVID-19 patient populations. Several clinical trials (namely RECOVERY and PANORAMIC) and observational studies of the two oral antivirals are ongoing, which will take into account the circulating Omicron variant and vaccination status of patients.^34-38^

The scientific community recognizes that logistics and distribution issues of oral antivirals should be adequately addressed by governments and the healthcare sector to facilitate drug initiation soon after symptom onset for maximal efficacy, and promote equitable access in the midst of limited supplies.^15,16,24^ For instance, a validated risk prediction tool or evidence-based scoring system can be developed to guide physicians to identify and prioritize COVID-19 patients who would most likely benefit from the use of oral antivirals.^16,26^ Besides, a number of research gaps remain in the evaluation of oral antiviral use, namely the safety of nirmatrelvir/ritonavir in children, pregnant or breastfeeding women, efficacy of oral antivirals in COVID-19 patients by serostatus at baseline, and risk of emergence of new viral variants attributed to genetic mutations induced by molnupiravir.^15,26,31^ Active pharmacovigilance programs are crucial to monitoring the long-term safety of oral antivirals, especially for the mutational risk and potential genotoxicity associated with molnupiravir use in light of conflicting experimental evidence.^39-42^ Moreover, given the high mutation rates of SARS-CoV-2 and selective pressure induced by the widespread use of antiviral monotherapy, concerns about the development of antiviral resistance have been raised.^15,26^ Further clinical studies are needed to examine the feasibility of combination therapy in accelerating the elimination of virus and minimizing drug resistance, for example, molnupiravir plus nirmatrelvir/ritonavir or favipiravir, or oral antivirals plus anti-SARS-CoV-2 monoclonal antibodies.^9,15,26,40^

Utilizing the public healthcare databases that encompass all reported cases of COVID-19 during the observation period, our study covers a non-selective patient population in the local region amid a pandemic wave of the Omicron variant. Alongside the introduction of both oral antivirals in the public healthcare system during this outbreak, their clinical effectiveness could be evaluated and compared in a real-world setting. Besides, consistent results were obtained from both the retrospective cohort and case-control analyses of our study, hence confirming the robustness of our findings. Nevertheless, a number of study limitations should be acknowledged. Firstly, residents at the RCHE were excluded from the current analysis because of substantial missing records, complex referral patterns between different levels and categories of treatment facilities and/or prolonged delays in oral antiviral prescription during the peak of this pandemic wave. Further studies are needed to examine the real-world safety and effectiveness of oral antivirals in specific healthcare settings, for instance, nursing homes and residential care facilities. Secondly, indication bias could not be eliminated in the prescription of oral antivirals, as reflected by the considerably older age and lower percentage of patients who had been fully vaccinated among oral antiviral users than matched controls at baseline. Indication bias might also be present in the clinical decision to prescribe molnupiravir versus nirmatrelvir/ritonavir, as the latter could be confounded by its significant drug-drug interactions. After matching, patient characteristics between oral antiviral and respective control groups were well balanced at baseline. Thirdly, some information biases might exist in the collection of data during the peak of this pandemic wave, such as the self-reporting of COVID-19 cases based on positive RAT with varying sensitivity. Lastly, there might have been an underreporting of COVID-19 cases during the study period, and the overwhelmed public healthcare system might have prevented some patients who would have been hospitalized from being admitted at the peak of the pandemic wave.

During a pandemic wave of SARS-CoV-2 Omicron variant, early initiation of molnupiravir or nirmatrelvir/ritonavir within five days of symptom onset among community-dwelling COVID-19 patients was associated with significant reduction of all-cause mortality risk by 39% and 75%, respectively, compared to not using any oral antivirals. Nirmatrelvir/ritonavir use was also associated with a reduced risk of COVID-19-related hospitalization by 31%, which was consistently observed across age and vaccination status. Further research is needed to confirm our results in specific patient populations and other healthcare settings.

## Supporting information

STROBE checklist

## Data Availability

The clinical outcome data were extracted from the Hospital Authority database in Hong Kong and vaccination records were extracted from the eSARS data provided by the Centre for Health Protection. Restrictions apply to the availability of these data, which were used under license for this study.

## Contributors

The study was designed by CKHW, GML and BJC. The underlying data was verified by CKHW, ICHA and EHYL, and data analyses were done by CKHW and ICHA. CKHW and KTKL wrote the first draft of the manuscript which was revised by GML and BJC. All authors interpreted data, provided critical review and revision of the text and approved the final version of the manuscript.

