## Supplementary files for "Real-world effectiveness of molnupiravir and nirmatrelvir/ritonavir against mortality, hospitalization, and in-hospital outcomes among community-dwelling, ambulatory COVID-19 patients during the BA.2.2 wave in Hong Kong: an observational study"

**Supplementary Material**

Supplementary Table 1. Baseline characteristics of non-hospitalized patients with COVID-19 in (a) outpatient molnupiravir and respective matched control groups, and (b) outpatient nirmatrelvir/ritonavir and respective matched control groups before 1:10 propensity score matching

Supplementary Table 2. Characteristics of case and control groups for outcomes of all-cause mortality, hospitalization, and composite in-hospital outcome after matching

Supplementary Table 3. Subgroup analyses of outcomes for (a) outpatient molnupiravir users versus matched controls, and (b) outpatient nirmatrelvir/ritonavir users versus matched controls by vaccination status and age groups

**Supplementary Table 1. Baseline characteristics of non-hospitalized patients with COVID-19 in (a) outpatient molnupiravir and respective matched control groups, and (b) outpatient nirmatrelvir/ritonavir and respective matched control groups before 1:10 propensity score matching**

| Baseline characteristics | Before 1:10 matching | | | | | | | | | |
|---|---|---|---|---|---|---|---|---|---|---|
| | Molnupiravir (n=5,257) | | Control (n=918,309) | | SMD | Nirmatrelvir/ritonavir (n=5,663) | | Control (n=918,309) | | SMD |
| | N / Mean | % / SD | N / Mean | % / SD | | N / Mean | % / SD | N / Mean | % / SD | |
| Age, years † | 75.0 | 12.6 | 49.2 | 16.7 | 1.54 | 70.4 | 11.9 | 49.2 | 16.7 | 1.27 |
| 18-40 | 88 | (1.7%) | 319,988 | (34.9%) | | 135 | (2.4%) | 319,988 | (34.9%) | |
| 40-65 | 925 | (17.6%) | 433,792 | (47.2%) | 1.70 | 1,594 | (28.2%) | 433,792 | (47.2%) | 1.36 |
| >65 | 4,244 | (80.7%) | 164,529 | (17.9%) | | 3,934 | (69.5%) | 164,529 | (17.9%) | |
| Sex | | | | | 0.03 | | | | | 0.00 |
| Male | 2,501 | (47.6%) | 423,790 | (46.1%) | | 2,609 | (46.1%) | 423,790 | (46.1%) | |
| Female | 2,756 | (52.4%) | 494,519 | (53.9%) | | 3,054 | (53.9%) | 494,519 | (53.9%) | |
| Pre-existing comorbidities | | | | | | | | | | |
| Charlson's Index † | 3.9 | 1.3 | 1.7 | 1.5 | 1.42 | 3.4 | 1.1 | 1.7 | 1.5 | 1.11 |
| 0-4 | 4,517 | (85.9%) | 912,655 | (99.4%) | | 5,373 | (94.9%) | 912,655 | (99.4%) | |
| 5-6 | 498 | (9.5%) | 3,724 | (0.4%) | 0.53 | 243 | (4.3%) | 3,724 | (0.4%) | 0.27 |
| 7-14 | 242 | (4.6%) | 1,930 | (0.2%) | | 47 | (0.8%) | 1,930 | (0.2%) | |
| Fully vaccinated* | 859 | (16.3%) | 501,302 | (54.6%) | 0.87 | 1,992 | (35.2%) | 501,302 | (54.6%) | 0.40 |

Notes: SD = standard deviation; SMD = standardized mean difference

* Fully vaccinated patients were defined as those with at least 2 doses of Comirnaty or 3 doses of CoronaVac.

† Age and Charlson Comorbidity Index are presented in mean ± SD.

**Supplementary Table 2. Characteristics of case and control groups for outcomes of all-cause mortality, hospitalization, and composite in-hospital outcome after matching**

| Characteristics | All-cause mortality | | | | Hospitalization | | | | Composite in-hospital outcome | | | |
|---|---|---|---|---|---|---|---|---|---|---|---|---|
| | Case (n=1,084) | | Control (n=10,454) | | Case (n=2,052) | | Control (n=19,099) | | Case (n=880) | | Control (n=8,438) | |
| | N / Mean | % / SD | N / Mean | % / SD | N / Mean | % / SD | N / Mean | % / SD | N / Mean | % / SD | N / Mean | % / SD |
| Age, years † | 80.0 | 13.5 | 80.7 | 11.4 | 55.1 | 20.3 | 54.6 | 17.7 | 74.5 | 15.7 | 74.9 | 14.2 |
| 18-40 | 18 | (1.7%) | 90 | (0.9%) | 654 | (31.9%) | 4,653 | (24.4%) | 37 | (4.2%) | 204 | (2.4%) |
| 40-65 | 137 | (12.6%) | 855 | (8.2%) | 695 | (33.9%) | 8,752 | (45.8%) | 190 | (21.6%) | 1,755 | (20.8%) |
| >65 | 929 | (85.7%) | 9,509 | (91.0%) | 703 | (34.3%) | 5,694 | (29.8%) | 653 | (74.2%) | 6,479 | (76.8%) |
| Sex | | | | | | | | | | | | |
| Male | 643 | (59.3%) | 6,128 | (58.6%) | 908 | (44.2%) | 8,504 | (44.5%) | 515 | (58.5%) | 4,813 | (57.0%) |
| Female | 441 | (40.7%) | 4,326 | (41.4%) | 1,144 | (55.8%) | 10,595 | (55.5%) | 365 | (41.5%) | 3,625 | (43.0%) |
| Pre-existing comorbidities | | | | | | | | | | | | |
| Charlson's Index †‡ | 5.1 | 2.0 | 4.9 | 1.8 | 2.3 | 2.1 | 2.2 | 1.6 | 5.5 | 2.1 | 5.1 | 2.1 |
| 1-4 | 540 | (49.8%) | 6,039 | (57.8%) | 1,738 | (84.7%) | 18,189 | (95.2%) | 264 | (30.0%) | 3,419 | (40.5%) |
| 5-6 | 312 | (28.8%) | 2,567 | (24.6%) | 265 | (12.9%) | 838 | (4.4%) | 377 | (42.8%) | 3,089 | (36.6%) |
| 7-14 | 232 | (21.4%) | 1,848 | (17.7%) | 49 | (2.4%) | 72 | (0.4%) | 239 | (27.2%) | 1,930 | (22.9%) |
| Full SARS-CoV-2 vaccination | 235 | (21.7%) | 2,047 | (19.6%) | 813 | (39.6%) | 8,458 | (44.3%) | 292 | (33.2%) | 2,886 | (34.2%) |

Notes: SD = standard deviation; SMD = standardized mean difference

* Fully vaccinated patients were defined as those with at least 2 doses of Comirnaty or 3 doses of CoronaVac.

† Age and Charlson Comorbidity Index are presented in mean ± SD.

**Supplementary Table 3. Subgroup analyses of outcomes for (a) outpatient molnupiravir users versus matched controls, and (b) outpatient nirmatrelvir/ritonavir users versus matched controls by vaccination status and age groups**

| Outcomes | Molnupiravir (N=4,875) | | | Control (N=48,409) | | | Molnupiravir vs Control | | | |
|---|---|---|---|---|---|---|---|---|---|---|
| | Crude incidence rate (Events / 100,000 person-days) | | | Crude incidence rate (Events / 100,000 person-days) | | | | | | |
| | Estimate | 95% CI | Person-days | Estimate | 95% CI | Person-days | HR† | 95% CI | P-value | $P_{int}$ |
| All-cause mortality | 24.2 | (17.8, 32.1) | 198,397 | 38.0 | (35.3, 40.7) | 2,033,869 | 0.61 | (0.46, 0.82) | <0.001 | |
| With full SARS-CoV-2 vaccination | 3.4 | (0.1, 18.7) | 29,724 | 9.1 | (5.7, 13.6) | 253,726 | NA | NA | NA | NA |
| Without full SARS-CoV-2 vaccination | 27.9 | (20.5, 37.1) | 168,673 | 42.1 | (39.1, 45.2) | 1,780,143 | 0.63 | (0.47, 0.85) | 0.002 | |
| Age ≤65 | 5.1 | (0.6, 18.2) | 39,601 | 6.5 | (4.2, 9.7) | 369,650 | 0.77 | (0.18, 3.30) | 0.726 | 0.730 |
| Age >65 | 29.0 | (21.2, 38.6) | 158,796 | 44.9 | (41.8, 48.3) | 1,664,219 | 0.61 | (0.45, 0.82) | 0.001 | |
| Hospitalization | 296.0 | (271.4, 322.3) | 179,704 | 269.2 | (261.7, 276.8) | 1,844,538 | 1.06 | (0.97, 1.16) | 0.191 | |
| With full SARS-CoV-2 vaccination | 115.1 | (79.2, 161.6) | 28,680 | 184.7 | (167.9, 202.7) | 239,306 | 0.62 | (0.44, 0.89) | 0.009 | 0.003 |
| Without full SARS-CoV-2 vaccination | 330.4 | (302.1, 360.7) | 151,024 | 281.8 | (273.6, 290.1) | 1,605,232 | 1.12 | (1.02, 1.23) | 0.013 | |
| Age ≤65 | 258.1 | (208.6, 315.9) | 36,418 | 168.9 | (155.6, 183.1) | 348,653 | 1.59 | (1.28, 1.99) | <0.001 | <0.001 |
| Age >65 | 305.7 | (277.7, 335.7) | 143,286 | 292.5 | (283.9, 301.3) | 1,495,885 | 0.99 | (0.89, 1.09) | 0.782 | |
| In-hospital disease progression | 31.9 | (24.5, 40.8) | 197,791 | 47.7 | (44.7, 50.8) | 2,024,497 | 0.64 | (0.50, 0.83) | <0.001 | |
| With full SARS-CoV-2 vaccination | 13.5 | (3.7, 34.6) | 29,598 | 18.6 | (13.7, 24.7) | 252,829 | 0.74 | (0.26, 2.05) | 0.557 | 0.845 |
| Without full SARS-CoV-2 vaccination | 35.1 | (26.7, 45.2) | 168,193 | 51.8 | (48.5, 55.3) | 1,771,668 | 0.65 | (0.50, 0.84) | 0.001 | |
| Age ≤65 | 20.3 | (8.8, 40.0) | 39,392 | 17.7 | (13.6, 22.5) | 368,061 | 1.17 | (0.56, 2.44) | 0.676 | 0.092 |
| Age >65 | 34.7 | (26.2, 45.2) | 158,399 | 54.3 | (50.8, 58.0) | 1,656,436 | 0.60 | (0.46, 0.79) | <0.001 | |
| In-hospital death | 23.2 | (17.0, 30.9) | 198,397 | 37.9 | (35.2, 40.6) | 2,033,869 | 0.59 | (0.44, 0.79) | <0.001 | |
| With full SARS-CoV-2 vaccination | 3.4 | (0.1, 18.7) | 29,724 | 9.1 | (5.7, 13.6) | 253,726 | NA | NA | NA | NA |
| Without full SARS-CoV-2 vaccination | 26.7 | (19.5, 35.7) | 168,673 | 42.0 | (39.0, 45.1) | 1,780,143 | 0.61 | (0.45, 0.82) | 0.001 | |
| Age ≤65 | 2.5 | (0.1, 14.1) | 39,601 | 6.5 | (4.2, 9.7) | 369,650 | NA | NA | NA | NA |
| Age >65 | 28.3 | (20.7, 37.9) | 158,796 | 44.8 | (41.7, 48.2) | 1,664,219 | 0.60 | (0.44, 0.81) | <0.001 | |
| Invasive mechanical ventilation | 4.0 | (1.7, 8.0) | 198,157 | 8.7 | (7.4, 10.0) | 2,030,226 | 0.45 | (0.22, 0.91) | 0.027 | |
| With full SARS-CoV-2 vaccination | 0.0 | NA | 29,724 | 4.7 | (2.4, 8.3) | 253,415 | NA | NA | NA | NA |

| | | | | | | | | | | |
|---|---|---|---|---|---|---|---|---|---|---|
| Without full SARS-CoV-2 vaccination | 4.7 | (2.1, 9.4) | 168,433 | 9.2 | (7.9, 10.8) | 1,776,811 | 0.50 | (0.24, 1.01) | 0.053 | |
| Age ≤65 | 7.6 | (1.6, 22.2) | 39,528 | 6.0 | (3.7, 9.0) | 368,913 | 1.26 | (0.38, 4.18) | 0.706 | 0.067 |
| Age >65 | 3.2 | (1.0, 7.4) | 158,629 | 9.3 | (7.9, 10.9) | 1,661,313 | 0.32 | (0.13, 0.79) | 0.013 | |
| Intensive care unit admission | 9.1 | (5.4, 14.4) | 197,829 | 11.7 | (10.2, 13.2) | 2,025,071 | 0.75 | (0.46, 1.21) | 0.234 | |
| With full SARS-CoV-2 vaccination | 10.1 | (2.1, 29.6) | 29,598 | 9.5 | (6.1, 14.1) | 252,833 | 1.04 | (0.31, 3.50) | 0.944 | 0.535 |
| Without full SARS-CoV-2 vaccination | 8.9 | (5.0, 14.7) | 168,231 | 12.0 | (10.4, 13.7) | 1,772,238 | 0.71 | (0.42, 1.20) | 0.203 | |
| Age ≤65 | 17.8 | (7.1, 36.6) | 39,392 | 11.1 | (8.0, 15.1) | 368,103 | 1.65 | (0.74, 3.68) | 0.221 | 0.031 |
| Age >65 | 6.9 | (3.5, 12.4) | 158,437 | 11.8 | (10.2, 13.5) | 1,656,968 | 0.55 | (0.30, 1.01) | 0.055 | |

| Outcomes | Nirmatrelvir/ritonavir (N=5,366) Crude incidence rate (Events / 100,000 person-days) | | | Control (N=53,289) Crude incidence rate (Events / 100,000 person-days) | | | Nirmatrelvir/ritonavir vs Control | | | |
|---|---|---|---|---|---|---|---|---|---|---|
| | Estimate | 95% CI | Person-days | Estimate | 95% CI | Person-days | HR† | 95% CI | P-value | $P_{int}$ |
| All-cause mortality | 5.2 | (2.5, 9.6) | 192,552 | 20.1 | (18.1, 22.1) | 1,969,245 | 0.25 | (0.13, 0.47) | <0.001 | |
| With full SARS-CoV-2 vaccination | 0.0 | NA | 62,573 | 3.3 | (2.0, 5.0) | 611,867 | NA | NA | NA | NA |
| Without full SARS-CoV-2 vaccination | 7.7 | (3.7, 14.1) | 129,979 | 27.6 | (24.9, 30.6) | 1,357,378 | 0.27 | (0.14, 0.50) | <0.001 | |
| Age ≤65 | 0.0 | NA | 58,491 | 2.6 | (1.4, 4.3) | 548,884 | NA | NA | NA | NA |
| Age >65 | 7.5 | (3.6, 13.7) | 134,061 | 26.8 | (24.2, 29.7) | 1,420,361 | 0.26 | (0.14, 0.49) | <0.001 | |
| Hospitalization | 126.2 | (110.6, 143.5) | 185,406 | 178.8 | (172.7, 184.9) | 1,850,613 | 0.69 | (0.60, 0.79) | <0.001 | |
| With full SARS-CoV-2 vaccination | 61.6 | (43.6, 84.5) | 61,696 | 92.0 | (84.4, 100.0) | 594,672 | 0.65 | (0.47, 0.91) | 0.012 | 0.838 |
| Without full SARS-CoV-2 vaccination | 158.4 | (137.0, 182.2) | 123,710 | 219.8 | (211.7, 228.2) | 1,255,941 | 0.70 | (0.60, 0.81) | <0.001 | |
| Age ≤65 | 60.8 | (42.4, 84.6) | 57,525 | 115.4 | (106.5, 125.0) | 529,321 | 0.56 | (0.40, 0.79) | <0.001 | 0.314 |
| Age >65 | 155.6 | (134.7, 178.8) | 127,881 | 204.1 | (196.5, 212.0) | 1,321,292 | 0.70 | (0.61, 0.81) | <0.001 | |
| In-hospital disease progression | 12.5 | (8.0, 18.6) | 192,078 | 25.8 | (23.6, 28.2) | 1,964,548 | 0.47 | (0.31, 0.71) | <0.001 | |
| With full SARS-CoV-2 vaccination | 3.2 | (0.4, 11.6) | 62,505 | 7.7 | (5.7, 10.2) | 610,946 | 0.42 | (0.10, 1.73) | 0.230 | 0.835 |
| Without full SARS-CoV-2 vaccination | 17.0 | (10.6, 25.7) | 129,573 | 34.0 | (30.9, 37.2) | 1,353,602 | 0.48 | (0.31, 0.73) | <0.001 | |
| Age ≤65 | 3.4 | (0.4, 12.4) | 58,436 | 8.4 | (6.1, 11.2) | 547,824 | 0.42 | (0.10, 1.75) | 0.236 | 0.916 |
| Age >65 | 16.5 | (10.3, 24.9) | 133,642 | 32.5 | (29.6, 35.7) | 1,416,724 | 0.47 | (0.31, 0.72) | <0.001 | |
| In-hospital death | 4.7 | (2.1, 8.9) | 192,552 | 20.0 | (18.1, 22.1) | 1,969,245 | 0.23 | (0.12, 0.44) | <0.001 | |

| | | | | | | | | | | |
|---|---|---|---|---|---|---|---|---|---|---|
| With full SARS-CoV-2 vaccination | 0.0 | NA | 62,573 | 3.3 | (2.0, 5.0) | 611,867 | NA | NA | NA | NA |
| Without full SARS-CoV-2 vaccination | 6.9 | (3.2, 13.1) | 129,979 | 27.6 | (24.8, 30.5) | 1,357,378 | 0.24 | (0.12, 0.47) | <0.001 | |
| Age ≤65 | 0.0 | NA | 58,491 | 2.6 | (1.4, 4.3) | 548,884 | NA | NA | NA | NA |
| Age >65 | 6.7 | (3.1, 12.7) | 134,061 | 26.8 | (24.1, 29.6) | 1,420,361 | 0.23 | (0.12, 0.45) | <0.001 | |
| Invasive mechanical ventilation | 2.1 | (0.6, 5.3) | 192,412 | 4.2 | (3.3, 5.2) | 1,967,711 | 0.48 | (0.18, 1.31) | 0.151 | |
| With full SARS-CoV-2 vaccination | 1.6 | (0.0, 8.9) | 62,539 | 1.6 | (0.8, 3.0) | 611,683 | NA | NA | NA | NA |
| Without full SARS-CoV-2 vaccination | 2.3 | (0.5, 6.8) | 129,873 | 5.3 | (4.2, 6.7) | 1,356,028 | 0.41 | (0.13, 1.31) | 0.133 | |
| Age ≤65 | 0.0 | NA | 58,491 | 2.4 | (1.3, 4.1) | 548,584 | NA | NA | NA | NA |
| Age >65 | 3.0 | (0.8, 7.6) | 133,921 | 4.9 | (3.8, 6.2) | 1,419,127 | 0.57 | (0.21, 1.55) | 0.269 | |
| Intensive care unit admission | 8.3 | (4.8, 13.5) | 192,099 | 6.6 | (5.5, 7.8) | 1,964,733 | 1.22 | (0.73, 2.05) | 0.450 | |
| With full SARS-CoV-2 vaccination | 3.2 | (0.4, 11.6) | 62,505 | 4.4 | (2.9, 6.4) | 610,965 | 0.69 | (0.16, 2.93) | 0.617 | 0.414 |
| Without full SARS-CoV-2 vaccination | 10.8 | (5.9, 18.1) | 129,594 | 7.5 | (6.1, 9.1) | 1,353,768 | 1.37 | (0.78, 2.39) | 0.271 | |
| Age ≤65 | 3.4 | (0.4, 12.4) | 58,436 | 5.3 | (3.5, 7.6) | 547,876 | 0.70 | (0.17, 2.92) | 0.621 | 0.401 |
| Age >65 | 10.5 | (5.7, 17.6) | 133,663 | 7.1 | (5.7, 8.6) | 1,416,857 | 1.35 | (0.77, 2.35) | 0.298 | |

Notes: HR = hazard ratio; CI = confidence interval; NA = not applicable; $P_{int}$ = P-value for interaction

† HR >1 (or <1) indicates oral antiviral users had higher (lower) risk of outcome compared to the matched control group.

HR were estimated only when the number of events in both groups were more than or equal to 2.
